# Protocol for Implementation and Evaluation of a Reserve-Stress-Rescue Pathway for High-Risk Preoperative Triage

**DOI:** 10.64898/2026.07.09.26357629

**Authors:** Inwoo Sohn, Tanush Singh, Zyad Carr

**Author notes:** **Corresponding Author** Zyad J. Carr, M.D., FASA. These authors contributed equally to this work (Inwoo Sohn, Tanush Singh). Senior author (Zyad Carr).

## Abstract

**Background:** High-risk preoperative triage remains fragmented: existing tools often estimate risk without identifying modifiable mechanisms or linking classification to postoperative monitoring, destination planning, and rescue resources. This protocol describes implementation and evaluation of a Reserve-Stress-Rescue (RSR Framework), pathway that operationalizes perioperative high risk as a mismatch among patient physiologic reserve, procedural stress, and system rescue capacity.

**Approach:** RSR is a proposed clinician-facing, modular scoring framework for adults undergoing major surgery, especially patients with frailty, multimorbidity, poor functional capacity, anemia or malnutrition, cardiopulmonary disease, or limited postoperative support. Each domain, Reserve, Stress, and Rescue, is scored from 0 to 4 and recorded as both a three-part profile and a total score from 0 to 12. Scores map to **Green**, **Amber**, **Red**, and **Crimson** triage bands that trigger escalating actions, including targeted optimization, multidisciplinary review, anesthesia and surgical planning, postoperative destination selection, monitoring intensity, and predefined escalation criteria.

**Validation Plan:** The initial phase of this study received an exemption determination from the Yale University Institutional Review Board on June 3, 2026, under IRB Protocol ID 2000042729, with exempt categories 2(ii) and 4(iii), including a waiver of HIPAA authorization for access to and use of protected health information as described in the approved protocol. Evaluation will proceed in stages, assessing feasibility, interrater reliability, completeness, acceptability, discrimination, calibration, and clinical utility. Key outcomes include postoperative complications, unplanned escalation of care, intensive care utilization, failure to rescue, mortality, length of stay, triage burden, low-yield testing cascades, and management-changing pathway activation.

**Conclusion:** The RSR pathway reframes high-risk status as a modifiable interaction between vulnerability, operative insult, and rescue capacity rather than a fixed patient label. If feasible and valid, RSR may standardize high-risk identification, align perioperative resources with anticipated physiology, improve communication, and support safer, actionable shared decision-making.

## Introduction

There remains a practical gap in rapid, holistic preoperative triage tools that classify high risk consistently and link classification to management-changing actions(1). For example, the American Society of Anesthesiologists Physical Status (ASA-PS) remains durable because it captures clinician synthesis of disease burden and physiologic reserve on a familiar ordinal scale associated with postoperative outcomes, but it is more useful for risk communication and adjustment than for identifying mechanisms or directing mitigation(2). Existing tools are either organ-specific (cardiac-centered constructs like MACE or RCRI risk), procedure-anchored, or data-intensive calculators that produce a probability estimate without clearly specifying downstream actions. Consequently, preoperative evaluation may fragment into parallel “clearance” pathways that generate labels, testing, and consultations without producing a shared plan for postoperative destination, monitoring intensity, surveillance, or rescue triggers(3).

High-risk status is defined by the combination of baseline vulnerability (age, multimorbidity, frailty, poor functional capacity, malnutrition/anemia), high-stress procedures (major abdominal, thoracic, vascular, complex oncology), and resource limitations. We propose a clinician-facing preoperative triage framework and scoring rubric that outlines an empirical validation pathway to operationalize perioperative “high risk” as a modifiable mismatch between physiologic reserve, surgical stress, and system rescue capacity. The rubric is designed to support reproducible classification, management-changing pathway activation, and resource-aligned postoperative planning. Because implementation is system-dependent, RSR is intended to be modular and locally adaptable, with staged validation of feasibility, reliability, predictive performance, calibration, and clinical utility, including effects on unplanned escalation, failure to rescue, triage burden, and low-yield testing cascades.

### Defining High-Risk

High risk is an interaction state, not a binary patient trait, and identifying a patient as high risk should guide mitigation rather than justify denying surgery(4). The goal of high-risk preoperative evaluation is to determine why risk is elevated, optimize modifiable vulnerabilities, align monitoring and rescue resources with anticipated physiology, and support shared decision-making (see **Table 1**). High-risk preoperative evaluation is most effective when it moves toward structured risk estimation, targeting testing that changes management or reveals modifiable risk factors using comorbidities, structured surveys (Duke Activity Status Index(5), Frailty Index(6)), and individualized physiological parameters (cardiopulmonary exercise testing(7, 8), biomarkers(9)). RSR does not rely on a universal predicted-probability threshold; instead, it uses pragmatic ordinal thresholds to standardize triage intensity and activate perioperative pathways intended to reduce postoperative complications.

**Table 1.** High-risk features for common adverse outcomes mapped to mechanism, preferred preoperative measurement, and actionable changes in perioperative management. Features are organized as pragmatic pathway triggers and framed around a mismatch model in which baseline vulnerability and limited physiologic reserve interact with surgical stress and system capacity. The Best Measurement column prioritizes tools that can be obtained preoperatively, and the Care Recommendations column emphasizes interventions that alter perioperative decision-making (monitoring intensity, destination planning, hemodynamic/ventilatory strategy, and consultation), rather than risk scoring alone. Abbreviations: ABG arterial blood gas; AKI acute kidney injury; ASA aortic stenosis; BNP B-type natriuretic peptide; CAD coronary artery disease; CBC complete blood count; CFS Clinical Frailty Scale; CKD chronic kidney disease; CO cardiac output; COPD chronic obstructive pulmonary disease; CPET cardiopulmonary exercise testing; DASI Duke Activity Status Index; ECG electrocardiogram; EF ejection fraction; EBL estimated blood loss; HF heart failure; ILD interstitial lung disease; MI myocardial infarction; MS mitral stenosis; NIV noninvasive ventilation; NT-proBNP N-terminal pro–B-type natriuretic peptide; OHS obesity hypoventilation syndrome; OSA obstructive sleep apnea; PFT pulmonary function tests; PH pulmonary hypertension; RV right ventricle/right ventricular; TSAT transferrin saturation; VBG venous blood gas; 6MWT six-minute walk test.

| High-risk feature (trigger) | Mechanism | Best measurement in preop workflow | What changes in care |
| --- | --- | --- | --- |
| Poor functional capacity | Low integrated cardiopulmonary reserve; limited ability to increase oxygen delivery under stress | DASI, stair-climb, 6MWT; CPET if decision-changing | Escalate monitoring/postop destination; CPET-informed optimization; tighter hemodynamic targets |
| Frailty / sarcopenia | Low physiologic reserve across organs; vulnerable to immobilization, delirium, infection, catabolism | CFS, gait speed, grip strength; structured clinical assessment | Trigger prehab, delirium prevention bundle; plan stepdown/ICU destination |
| Anemia / iron deficiency | Reduced oxygen carrying capacity; worsens supply-demand mismatch; increases transfusion exposure | CBC; ferritin/TSAT when time allows | Preop iron/transfusion strategy; blood conservation; vigilance for ischemia/AKI |
| Malnutrition / hypoalbuminemia | Reduced stress tolerance and immune reserve; associated with poor wound healing and complications | Weight loss history; nutrition screen; albumin as severity marker | Nutrition optimization; postop feeding plan; consider deferral for severe elective cases |
| Heart failure (reduced EF or evidence of congestion) | Limited ability to augment cardiac output; fluid shifts and hypotension precipitate hypoperfusion and congestion-driven AKI | Symptoms/exam; BNP/NT-proBNP; echo; volume status | Individualized fluid/vasoactive strategy; consider invasive monitoring; planned ICU/stepdown |
| CAD / prior MI / inducible ischemia | Limited coronary flow reserve; vulnerable to tachycardia, hypotension, anemia | History; ECG; BNP/NT-proBNP; stress testing only if management-changing | Tight HR/BP control; avoid prolonged hypotension; telemetry/troponin surveillance as indicated |
| Valvular disease (AS/MS) or pulmonary hypertension | Fixed-flow states or high RV afterload; decompensate with vasodilation, hypoxia, hypercarbia | Echo; symptoms; BNP/NT-proBNP when helpful | Anesthetic plan around RV/LV physiology; avoid hypoxia/hypercarbia; low threshold for ICU |
| Pulmonary disease with limited ventilatory reserve (COPD/ILD/OSA/OHS) | Higher risk of ventilatory failure, gas-exchange impairment, atelectasis; spillover into cardiac/renal stress | History; PFTs when helpful; ABG/VBG if hypercapnia suspected | Lung-protective ventilation; postop NIV planning; aggressive pulmonary toilet and early mobilization |
| CKD / prior AKI / albuminuria | Reduced renal reserve; susceptible to hypotension, congestion, inflammation, nephrotoxins | Creatinine/eGFR; urine albumin/ACR; prior AKI history | Avoid nephrotoxins; tighter MAP goals; early postop labs; diuresis/congestion plan |
| High-stress procedure (major abdominal/thoracic/vascular/complex oncology) | Large inflammatory load, fluid shifts, bleeding risk, prolonged duration drives global mismatch | Procedure category; expected duration/EBL; surgeon input | Default pathway activation; plan monitoring/lines; ICU/stepdown destination planning |
| Limited system capacity / poor rescue | Risk amplified when escalation is delayed or monitoring is insufficient | Institutional resources and local constraints | Pre-plan destination; standardize triggers; ensure rescue resources or adjust plan |

Clinically meaningful outcomes, including mortality and disability, rarely arise from a single isolated complication. Adverse events span myocardial injury(10), respiratory failure(11), acute kidney injury(12), delirium(13), infection(14), venous thromboembolism, prolonged ileus, pain syndromes, and functional decline(15). The patient-centered consequences are often similar: loss of physiologic reserve, prolonged recovery, reduced independence, and poorer quality of life(16–19). From this perspective, high-risk preoperative evaluation should identify patients likely to experience postoperative deconditioning, organ injury, or failure to return to baseline(20, 21). This reframing helps reconcile why traditional labels (e.g., vascular/high-stress surgery, multimorbidity, frailty, or abnormal biomarkers) can all correctly signal “high-risk” while still being heterogeneous and inconsistently actionable(22).

In current practice, high-risk status is often defined by predicted MACE thresholds, elevated RCRI, high-stress or emergent surgery, poor functional capacity, frailty, multimorbidity, or elevated biomarkers such as natriuretic peptides or high-sensitivity troponin(23, 24). These approaches are useful for identifying at-risk patients but often emphasize risk stratification without specifying management-changing actions. For this protocol, we define high risk as a state in which surgical stress is likely to exceed the patient’s functional reserve and/or the system’s capacity to detect and rescue deterioration. The definition focuses on clinically meaningful outcomes, including readmission, loss of independence, prolonged functional limitation, and major cardiac, pulmonary, or renal complications, while supporting reproducible and efficient triage.

This lack of a unifying triage layer matters because perioperative harm often reflects a trajectory of physiologic decompensation and delayed recognition, not a single event. Existing tools rarely integrate the three determinants of that trajectory: patient reserve, surgical stress, and rescue capacity. As a result, patients with similar comorbidities may face different risks depending on procedural intensity and postoperative environment, yet risk is often treated as a fixed patient attribute. A workflow-integrated triage tool should standardize team communication, reduce low-yield testing cascades, and map risk classification directly to bundled perioperative pathways.

### Proposed Reserve-Stress-Rescue (RSR) framework

Identifying high-risk patients before surgery is challenging because perioperative harm is rarely explained by a single diagnosis or test. Instead, outcomes reflect a dynamic mismatch among physiologic reserve, operative stress, and the system’s capacity to detect and rescue early deterioration(25). Because perioperative risk is common, time-sensitive, and shaped by patient, procedural, and system factors, clinicians need rapid triage tools that can be applied consistently at the point of care. Conventional assessment often relies on clinician gestalt or low-yield testing cascades, producing risk labels that do not reliably change management. Unlike probabilistic calculators, the RSR framework is designed to identify modifiable mismatches among physiologic demand, patient reserve, and system response capacity.

This pragmatically frames “high-risk” as a *mismatch* rather than a fixed patient label(26). Complications occur when surgical **Stress** exceeds physiologic **Reserve** in a care environment with insufficient **Rescue** capacity. RSR therefore shifts preoperative evaluation from “clearance” toward triage and mitigation: optimizing vulnerabilities when time allows, reducing operative and anesthetic stress where feasible, and aligning postoperative destination, monitoring, surveillance, and escalation triggers with anticipated physiology. As a three-domain tool, RSR provides both a total risk signal and an interpretable **R-S-Re** profile that maps directly to pathway activation and limits low-yield testing by tying diagnostics to pre-specified management decisions (see **Table 2**).

**Table 2.**
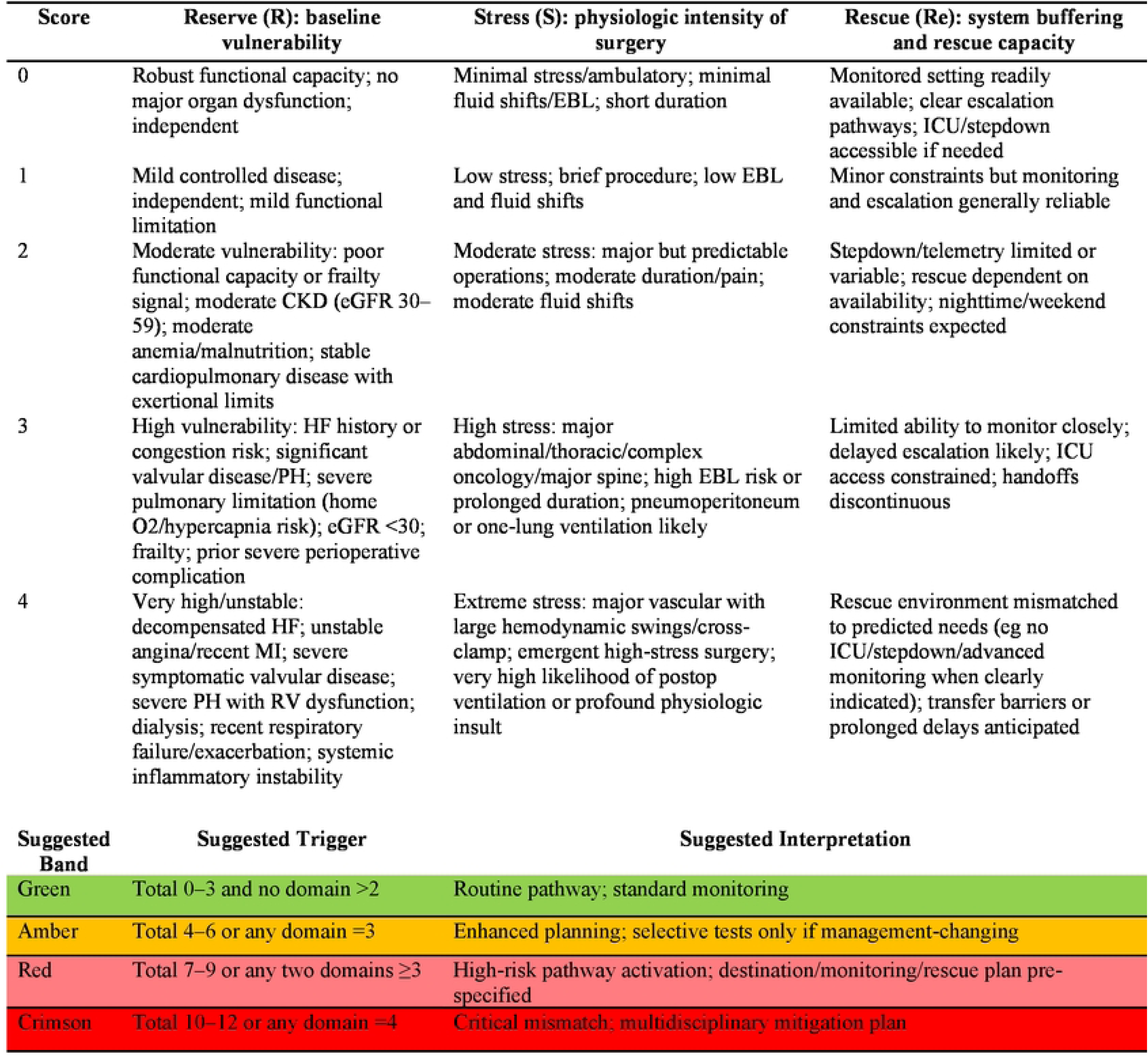
Clinician-driven Reserve–Stress–Rescue (RSR) assessment tool for preoperative triage. The RSR score operationalizes perioperative risk as a three-vector mismatch between Reserve (baseline physiologic vulnerability), Stress (physiologic intensity of the planned operation), and Rescue (system capacity to monitor, escalate, and intervene). Each domain is scored from 0 (lowest) to 4 (highest) by selecting the tier that best fits the patient, procedure, and care environment; the score may be recorded as both a profile (R,S,Re) to identify the dominant driver of risk and a total score (0–12) to support pathway activation. The lower panel provides suggested triage bands (Green, Amber, Red, Crimson) that map total score and/or domain thresholds to increasing levels of planning intensity, monitoring, postoperative destination planning, and rescue resource alignment. This framework is intended to trigger risk mitigation and system alignment, not to deny surgery. Abbreviations: CKD chronic kidney disease; EBL estimated blood loss; eGFR estimated glomerular filtration rate; HF heart failure; ICU intensive care unit; MI myocardial infarction; PH pulmonary hypertension; RV right ventricle/right ventricular.

#### Vector A

##### Baseline Vulnerability (physiologic reserve)

Vector A captures baseline physiologic reserve, or the patient’s ability to tolerate perioperative stress and recover, prioritizing functional capacity, frailty, cardiopulmonary disease burden, nutritional status, anemia. Functional limitation and frailty capture multi-organ reserve loss and frequently signal lack of physiological reserve(27, 28). Similarly, anemia and malnutrition are comorbidities that directly modify oxygen delivery, inflammatory tolerance, and wound healing. Cardiac vulnerability, framed by coronary artery disease, heart failure, pulmonary hypertension, valvular disease, conduction disease, and limited chronotropic reserve, suggest oxygen delivery limitations under surgical stress. Pulmonary vulnerability is focused on chronic baseline hypercapnia or hypoxemia, or prior respiratory failure, where the key unifier is limited ventilatory reserve. Renal vulnerability is focused on CKD, albuminuria, prior AKI, nephrotoxic exposure, propensity for postoperative volume overload and baseline poor renal perfusion. The unifier is the ability to tolerate hemodynamic and inflammatory insults without loss of filtration. Global vulnerability modifiers may include immunocompetency, cognitive impairment, multimorbidity burden, and polypharmacy.

#### Vector B

##### Surgical Stress

Vector B defines the magnitude and character of the surgical stress imposed by a procedure. Rather than relying on procedural labels alone, it considers the anticipated burden of tissue injury, blood loss, fluid redistribution, ventilatory challenge, pain, and hemodynamic volatility. This allows the framework to classify operations according to the physiologic load they impose, which is the relevant exposure for perioperative mismatch. Surgical stress is determined by the combination of risk for: 1) major fluid shifts and risk of bleeding, 2) inflammatory load and tissue trauma, 3) operative duration and positioning constraints, 4) probability of aggregate hypotension and vasopressor exposure, 5) pain burden driving sympathetic activation and respiratory splinting, 6) need for special physiological exposures (one-lung ventilation, pneumoperitoneum, aortic cross-clamping, extracorporeal membrane oxygenation). The unifier is the likelihood of the procedure forcing prolonged periods of supply-demand mismatch related to Vector A. Stress assignment should be informed by predefined procedural categories and expected physiologic exposures to support reproducibility.

#### Vector C

##### System buffering capacity

Vector C represents rescue capacity, or the system’s ability to recognize and respond to perioperative deterioration before it becomes catastrophic. It includes monitoring, staffing, escalation pathways, ICU (Intensive Care Unit) availability, and the reliability of postoperative surveillance. This vector explains why the same patient undergoing the same operation may be appropriately classified differently across care settings and is a core determinant of whether early physiologic drift becomes a treatable deviation or an irreversible complication. Buffering capacity includes: 1) immediate access to ICU/stepdown with optimal staffing ratios; 2) invasive monitoring capability and clinician expertise, 3) availability of biomarker surveillance, availability of postoperative respiratory support (e.g. high flow nasal cannula or noninvasive positive-pressure ventilation) and 4) early mobilization resources, hemodynamic protocols, and blood product logistics. The key metric is failure to rescue, as the same patient and same surgery may be “High Risk” at Hospital A and “Moderate Risk” at Hospital B. This mismatch cascade explains why adverse outcomes often cluster when an initial physiologic stressor produces supply-demand mismatch, prompts potentially harmful compensation, causes second-hit injury, and is compounded by failure to rescue.

#### RSR Framework Interpretation

RSR interpretation should distinguish the total score from the underlying domain profile. Although the total RSR score provides a pragmatic signal of overall triage intensity, the framework’s conceptual value lies in identifying the specific *mismatch* among physiologic reserve, surgical stress, and rescue capacity. Therefore, each patient should be reported using both an aggregate score and a domain profile, such as R3-S2-Re1, so clinicians can see whether the dominant concern is baseline vulnerability, operative physiologic load, or inadequate surveillance/escalation capacity. Critical mismatch rules, such an isolated domain score of 4 or hazardous combinations across domains, should be explicitly defined because they may justify escalation even when the additive total score alone appears moderate.

### Potential Implications of the RSR Framework

#### Avoidance of Preoperative Testing Cascades

Preoperative testing should be guided by whether it changes management, not simply by whether it refines risk. A preoperative test is justified when it is likely to alter one or more of the following: the decision to proceed or defer, the surgical approach, the anesthetic/hemodynamic plan, the postoperative destination, or the need for preoperative optimization/consultation. The central question is not whether a test refines risk numerically, but whether it meaningfully changes what the team will do. In contrast, a test contributes to a *testing cascade* when it refines risk numerically but does not plausibly change the perioperative plan or when it produces incidental findings that generate downstream interventions with unclear benefit in the perioperative timeframe.

Under the three-vector model, the most valuable tests are those that clarify *reserve* (Vector A) in a way that meaningfully reclassifies risk or suggests actionable optimization, those that clarify *stress* exposure (Vector B) to support pathway activation and resource allocation, and those that clarify *system needs* (Vector C) by defining monitoring and rescue requirements.

#### Integration of RSR with evidence-based pathways that can improve postoperative complication rates

Within the RSR framework, bundles are most effective when explicitly matched to the dominant vulnerability domain (Reserve), anticipated physiologic demand (Stress), or system reliability gap (Rescue), rather than applied generically to all high-risk patients. The RSR framework favors bundled, protocolized pathways (rather than isolated “one-off” interventions), with each component providing clear physiologic or behavioral benefit and the bundle itself is designed for high-fidelity implementation through standardization, defaults, and team-based execution(29, 30).

Outcomes improve most reliably when “risk” activates focused pathways that standardize optimization, surveillance, and early treatment(31). For myocardial injury/MINS and cardiac risk, major perioperative society guidelines endorse structured risk assessment and targeted testing only when it changes management, with postoperative troponin surveillance in higher-risk patients increasingly supported. The MANAGE trial reduced major vascular complications, supporting the concept that surveillance coupled to an actionable pathway can improve postoperative cardiovascular outcomes(32). For heart failure risk, evidence-supported bundles emphasize congestion assessment, conservative fluid strategy, early recognition of decompensation, and continuity of indicated therapies when feasible(9, 33–35). For respiratory failure risk, bundles centered on lung-protective ventilation and postoperative pulmonary complications prevention have randomized support(36). For AKI risk, KDIGO provides a widely adopted prevention bundle approach (avoidance of nephrotoxins, optimizing volume/hemodynamics, creatinine monitoring) and a KDIGO-based hemodynamic optimization bundle reduced AKI in a cardiac surgery population(37, 38).

#### Practical Application

A triage score can reliably flag risk but cannot fully capture the clinical nuance that determines the right response. Preserving clinician autonomy is essential because perioperative risk is contextual and time-dependent. Two patients with the same RSR profile may warrant different bundled pathways based on factors the rubric intentionally does not overfit to, such as trajectory and reversibility of disease, competing risks, patient goals, procedural alternatives, and local resource realities. If pathway activation is overly prescriptive, clinicians quickly experience it as “checkbox medicine”, leading to workarounds, poor fidelity, and erosion of trust in the tool. Conversely, for high-risk patients, risk communication and aligning treatment intensity with patient values are central.

RSR execution directly leads to structured perioperative decision making. For example, a Green tier (0-3, no domain >2) triggers a cost-effective standard pathway, Amber (4-6 or any domain =3) triggers Enhanced Planning Pathway, Red (7-9 or any two domains >3) triggers a high-risk bundle set, while Crimson (10-12 or any domain =4) results in a critical mismatch bundle set which includes multidisciplinary review and destination planning. These profile logics could be further refined based on the domain, where if the reserve domain score is the highest, implying high frailty or poor cardiopulmonary reserve, bundles are selected that increase physiologic buffer and prevent predictable perioperative organ injury. If surgical stress is highest, the operation is the driver, and bundles that reduce intraoperative stress are selected. If rescue is highest, the system is the limiting factor, and bundles are selected that increase surveillance and escalation capacity (destination, monitoring, and explicit triggers). By doing so, this turns “risk scoring” into an exercise of which domain is most modifiable right now. Secondly, in addition to base bundles that come with specific band scoring, domain bundles can be added depending on the profile.

#### Reserve bundle (R ≥3)

The Reserve bundle is the “optimize vulnerability” package, aimed at increasing tolerance to a given surgical stress. It prioritizes anemia management, volume and congestion planning, medication reconciliation with hemodynamic intent, targeted nutrition or prehabilitation when feasible, and postoperative surveillance for myocardial injury or AKI when indicated. The goal is to replace diffuse consult cascades with a small set of high-yield actions. For example, selecting the Reserve bundle could trigger a single order set containing core optimization labs, prompts indicating which medications to hold or continue, and default postoperative surveillance aligned to risk. Prehabilitation sits primarily within this domain because it seeks to increase physiologic buffer before surgery through improvements in functional capacity, nutrition, anemia, strength, and symptom control.

#### Stress bundle (S ≥3)

The Stress bundle is the “reduce physiologic load” package, focused on limiting the burden imposed by the operation and anesthetic. It standardizes blood conservation measures, hemodynamic targets, analgesic planning to reduce sympathetic stress, ventilation strategies for high-risk pulmonary mechanics, and early mobilization or feeding goals. It may also include a structured intraoperative handoff to improve continuity into PACU or ICU. In practice, bundle activation could generate a multidisciplinary co-signed plan template and perioperative order set centered on blood management, temperature management, and explicit physiologic targets.

#### Rescue bundle (Re ≥3)

The Rescue bundle is the “anticipate deterioration and escalate early” package. Because failure to rescue reflects not only complication risk but also the system’s ability to detect and respond to treatable deterioration, this domain is the most immediately actionable. It standardizes postoperative destination, monitoring intensity, escalation criteria, and early surveillance in the first 6–24 hours, including vital sign frequency, selected laboratory monitoring, and urine output expectations where relevant. For example, activating the Rescue bundle could place destination and monitoring orders, and attach standardized escalation algorithms to nursing workflows.

Pathways selection should be clinician-driven but constrained. The RSR framework works best when clinicians retain judgement, but the system is geared toward consistency, a concept well validated in the literature. A simple rule set that preserves choice would specify that the band determines minimum bundle intensity, where clinicians shouldn’t de-escalate without documentation. Secondly, the highest domain determines which add-on bundles are recommended by default. Clinicians should retain the ability to uncheck recommended bundles but must justify their reason from a short list (e.g. palliative goals, procedure cancelled, monitoring not available, competing diagnosis).

## Materials and Methods

### Aim, Design and Setting

This staged, single-center implementation and validation study will evaluate the Reserve-Stress-Rescue framework in the preoperative surgical triage setting by assessing four linked aims: implementation feasibility and completeness, interrater reliability of domain scoring and triage-band assignment, predictive validity and calibration for postoperative escalation and complications, and clinical utility in activating management-changing perioperative pathways (see **Table 3**). The initial phase of this study received an exemption determination from the Yale University Institutional Review Board on June 3, 2026, under IRB Protocol ID 2000042729, with exempt categories 2(ii) and 4(iii), including a waiver of HIPAA authorization for access to and use of protected health information as described in the approved protocol.

**Table 3.**
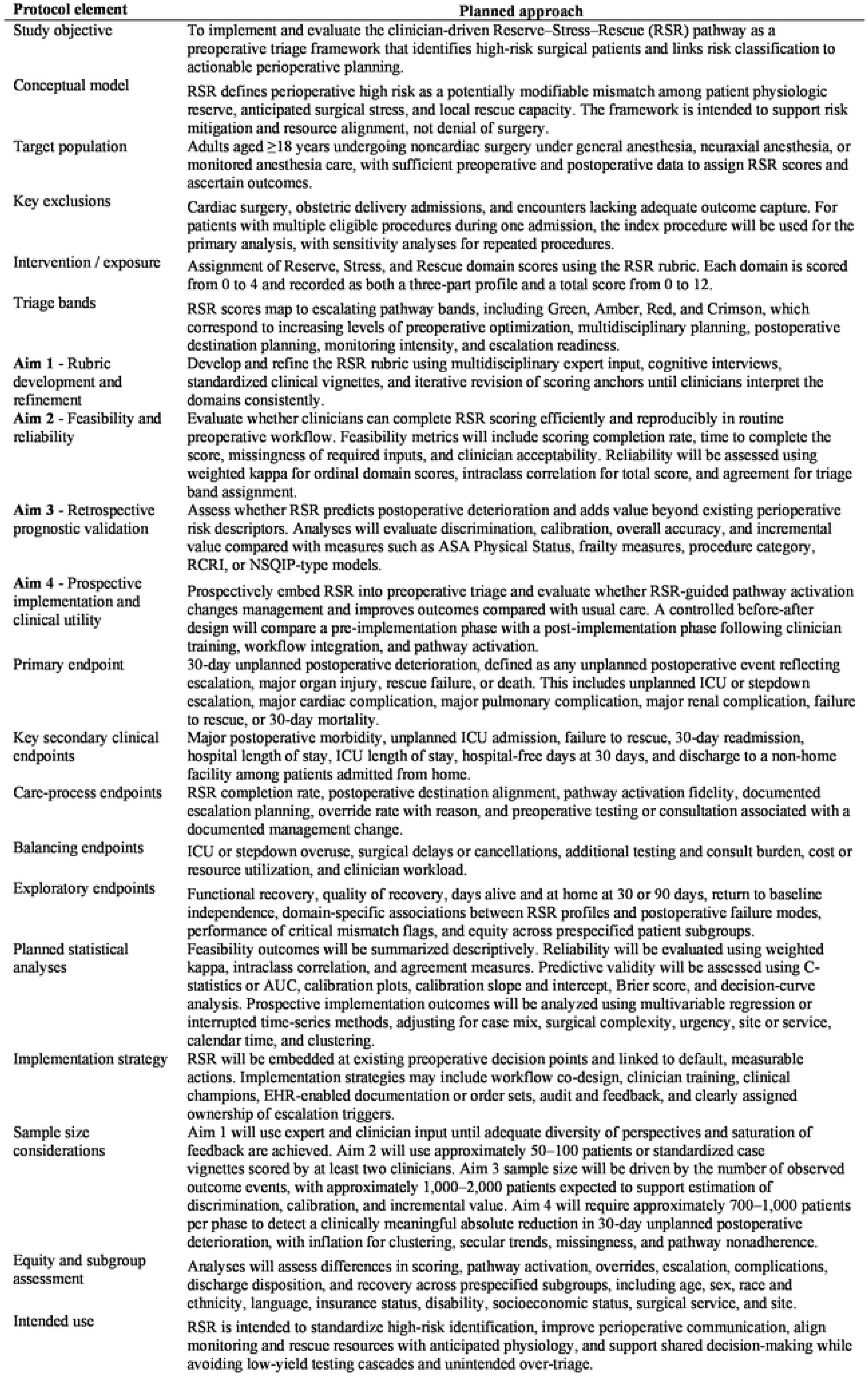
Summarizes the proposed staged protocol for developing, validating, and implementing the Reserve–Stress–Rescue pathway as a preoperative triage framework linking risk classification to actionable perioperative planning and outcome evaluation. RSR - Reserve–Stress–Rescue; ASA - American Society of Anesthesiologists Physical Status; RCRI - Revised Cardiac Risk Index; NSQIP - National Surgical Quality Improvement Program; ICU - intensive care unit; AUC - area under the receiver operating characteristic curve; EHR - electronic health record.

| Summary of the proposed staged protocol for development, validation and implementation of the Reserve-Stress-Rescue Pathway. |  |
| --- | --- |
| Protocol element | Planned approach |
| Study objective | To implement and evaluate the clinician-driven Reserve–Stress–Rescue (RSR) pathway as a preoperative triage framework that identifies high-risk surgical patients and links risk classification to actionable perioperative planning. |
| Conceptual model | RSR defines perioperative high risk as a potentially modifiable mismatch among patient physiologic reserve, anticipated surgical stress, and local rescue capacity. The framework is intended to support risk mitigation and resource alignment, not denial of surgery. |
| Target population | Adults aged ≥18 years undergoing noncardiac surgery under general anesthesia, neuraxial anesthesia, or monitored anesthesia care, with sufficient preoperative and postoperative data to assign RSR scores and ascertain outcomes. |
| Key exclusions | Cardiac surgery, obstetric delivery admissions, and encounters lacking adequate outcome capture. For patients with multiple eligible procedures during one admission, the index procedure will be used for the primary analysis, with sensitivity analyses for repeated procedures. |
| Intervention / exposure | Assignment of Reserve, Stress, and Rescue domain scores using the RSR rubric. Each domain is scored from 0 to 4 and recorded as both a three-part profile and a total score from 0 to 12. |
| Triage bands | RSR scores map to escalating pathway bands, including Green, Amber, Red, and Crimson, which correspond to increasing levels of preoperative optimization, multidisciplinary planning, postoperative destination planning, monitoring intensity, and escalation readiness. |
| **Aim 1** - Rubric development and refinement | Develop and refine the RSR rubric using multidisciplinary expert input, cognitive interviews, standardized clinical vignettes, and iterative revision of scoring anchors until clinicians interpret the domains consistently. |
| **Aim 2** - Feasibility and reliability | Evaluate whether clinicians can complete RSR scoring efficiently and reproducibly in routine preoperative workflow. Feasibility metrics will include scoring completion rate, time to complete the score, missingness of required inputs, and clinician acceptability. Reliability will be assessed using weighted kappa for ordinal domain scores, intraclass correlation for total score, and agreement for triage band assignment. |
| **Aim 3** - Retrospective prognostic validation | Assess whether RSR predicts postoperative deterioration and adds value beyond existing perioperative risk descriptors. Analyses will evaluate discrimination, calibration, overall accuracy, and incremental value compared with measures such as ASA Physical Status, frailty measures, procedure category, RCRI, or NSQIP-type models. |
| **Aim 4** - Prospective implementation and clinical utility | Prospectively embed RSR into preoperative triage and evaluate whether RSR-guided pathway activation changes management and improves outcomes compared with usual care. A controlled before-after design will compare a pre-implementation phase with a post-implementation phase following clinician training, workflow integration, and pathway activation. |
| Primary endpoint | 30-day unplanned postoperative deterioration, defined as any unplanned postoperative event reflecting escalation, major organ injury, rescue failure, or death. This includes unplanned ICU or stepdown escalation, major cardiac complication, major pulmonary complication, major renal complication, failure to rescue, or 30-day mortality. |
| Key secondary clinical endpoints | Major postoperative morbidity, unplanned ICU admission, failure to rescue, 30-day readmission, hospital length of stay, ICU length of stay, hospital-free days at 30 days, and discharge to a non-home facility among patients admitted from home. |
| Care-process endpoints | RSR completion rate, postoperative destination alignment, pathway activation fidelity, documented escalation planning, override rate with reason, and preoperative testing or consultation associated with a documented management change. |
| Balancing endpoints | ICU or stepdown overuse, surgical delays or cancellations, additional testing and consult burden, cost or resource utilization, and clinician workload. |
| Exploratory endpoints | Functional recovery, quality of recovery, days alive and at home at 30 or 90 days, return to baseline independence, domain-specific associations between RSR profiles and postoperative failure modes, performance of critical mismatch flags, and equity across prespecified patient subgroups. |
| Planned statistical analyses | Feasibility outcomes will be summarized descriptively. Reliability will be evaluated using weighted kappa, intraclass correlation, and agreement measures. Predictive validity will be assessed using C-statistics or AUC, calibration plots, calibration slope and intercept, Brier score, and decision-curve analysis. Prospective implementation outcomes will be analyzed using multivariable regression or interrupted time-series methods, adjusting for case mix, surgical complexity, urgency, site or service, calendar time, and clustering. |
| Implementation strategy | RSR will be embedded at existing preoperative decision points and linked to default, measurable actions. Implementation strategies may include workflow co-design, clinician training, clinical champions, EHR-enabled documentation or order sets, audit and feedback, and clearly assigned ownership of escalation triggers. |
| Sample size considerations | Aim 1 will use expert and clinician input until adequate diversity of perspectives and saturation of feedback are achieved. Aim 2 will use approximately 50–100 patients or standardized case vignettes scored by at least two clinicians. Aim 3 sample size will be driven by the number of observed outcome events, with approximately 1,000–2,000 patients expected to support estimation of discrimination, calibration, and incremental value. Aim 4 will require approximately 700–1,000 patients per phase to detect a clinically meaningful absolute reduction in 30-day unplanned postoperative deterioration, with inflation for clustering, secular trends, missingness, and pathway nonadherence. |
| Equity and subgroup assessment | Analyses will assess differences in scoring, pathway activation, overrides, escalation, complications, discharge disposition, and recovery across prespecified subgroups, including age, sex, race and ethnicity, language, insurance status, disability, socioeconomic status, surgical service, and site. |
| Intended use | RSR is intended to standardize high-risk identification, improve perioperative communication, align monitoring and rescue resources with anticipated physiology, and support shared decision-making while avoiding low-yield testing cascades and unintended over-triage. |

### Evaluation Objectives

The **first aim** will be to develop and refine the RSR rubric using a broad diaspora of physician and non-physician preoperative clinic settings. Using Delphi, or modified Delphi panel with perioperative stakeholders and cognitive interviews with intended users after informed consent. Pilot scoring of standardized vignettes will be followed by revision of anchors until clinicians interpret each score consistently (see **Supplementary File 1**).

The **second aim** will establish feasibility and inter-rater reliability, testing whether clinicians can assign Reserve, Stress, and Rescue scores consistently and efficiently using the **Table 2** rubric. Inter-rater reliability for each domain and composite profile will be quantified, and the rubric refined if agreement is suboptimal. Metrics will include time to complete the score, missingness, clinician acceptability and inter-rater agreement for each domain and total band. This will be measured using completion time, missingness, acceptability, weighted kappa, intraclass correlation (ICC) coefficient, and agreement for triage band.

**Aim 3** will evaluate predictive performance in a retrospective cohort of adult noncardiac surgeries by assessing discrimination and calibration for adverse outcomes (e.g., disposition, postoperative complications) to determine if RSR predicts outcomes and adds value beyond existing tools. This will be performed by benchmarking incremental value vs. common comparators such as ASA Class, procedure category, and established risk indices. We will assess this using discrimination (C-statistic/AUC), calibration plots, Brier score, and clinical utility (decision-curve analysis).

If successful, **Aim 4** will evaluate whether RSR-guided care improves outcomes by prospectively testing clinical utility. This will be achieved by embedding RSR scoring into preoperative triage, monitoring intensity, selective testing tied to pre-specified actions, and standardized postoperative surveillance. This will use a controlled before-after implementation study comparing usual preoperative triage with RSR-guided pathway activation. During the pre-implementation phase, eligible adult noncardiac surgery patients would receive standard preoperative evaluation, with outcomes and process measures collected prospectively but without visible RSR scoring. After clinician training, EHR integration, and activation of Reserve, Stress, and Rescue pathway bundles, the post-implementation phase would apply RSR scoring at the preoperative decision point and link tier/domain profiles to default management actions. The primary impact analysis would compare rates of unplanned postoperative escalation, failure to rescue, and major postoperative morbidity before versus after implementation, while secondary analyses would assess pathway fidelity, postoperative destination alignment, testing and subspecialty consult utilization, discharge to non-home facility, readmission, ICU use, surgical delays, and equity across patient subgroups. To strengthen causal inference, the analysis will adjust for case mix, surgical complexity, baseline secular trends, and clustering by service or site, with balancing measures used to detect unintended over-triage or resource strain.

### Sample Size Considerations

Because this is a staged implementation and validation protocol, sample size will be tailored to each evaluation aim. For the initial feasibility phase, we anticipate enrolling approximately 50-100 participants, which would allow estimation of pathway completion, missingness, adherence, and acceptability with approximately ±7.5–10% precision for expected rates of 80–90%. Interrater reliability will be assessed in a subset of at least 50 independently double-rated cases, with 75–100 cases preferred to evaluate agreement across RSR domain scores, total score, triage band assignment, and critical mismatch rules. For predictive validation, assuming a postoperative complication or unplanned escalation event rate of 15–25% and an anticipated C-statistic of approximately 0.70, approximately 300–500 patients would provide early estimates of discrimination with moderate precision, whereas 750–1,500 patients would be preferred for more precise discrimination, calibration, and subgroup analyses. A subsequent comparative effectiveness or clinical utility phase would require a larger sample, likely 1,000–3,000 patients, to detect plausible absolute reductions of 3–7% in clinically meaningful postoperative outcomes.

#### Endpoints

The primary endpoint will be 30-day unplanned postoperative deterioration, defined as unplanned ICU or stepdown escalation, major postoperative cardiac, pulmonary, or renal complication, failure to rescue, or death. Key secondary clinical endpoints will include major postoperative morbidity, unplanned ICU admission, failure to rescue, 30-day readmission, hospital and ICU length of stay, hospital-free days at 30 days, and discharge to a non-home facility among patients admitted from home. Care-process endpoints will include RSR completion rate, postoperative destination alignment, pathway activation fidelity, documented escalation planning, override rate with reason, and preoperative testing or consultation associated with a documented management change. Balancing endpoints will include ICU or stepdown overuse, surgical delays or cancellations, additional testing and consult burden, cost/resource utilization, and clinician workload.

Exploratory analyses will assess functional recovery, quality of recovery, days alive and at home (30 or 90 days), return to baseline independence, domain-specific associations between RSR profiles and postoperative failure modes, and equity across prespecified patient subgroups.

Study findings will be disseminated through peer-reviewed publication, conference presentation, and sharing of non-identifiable protocol materials through appropriate repositories. Any substantive protocol amendment or decision to terminate the study will be documented by the investigative team and submitted to the Yale Institutional Review Board for review and approval or confirmation of continued exempt status before implementation, except when immediate action is necessary to protect participant safety or privacy.

## Discussion

### Implementation and Safety Considerations

Even a strong construct won’t improve outcomes unless it can be executed with high adoption, fidelity, and sustainment in real workflows. In practical terms, RSR needs to be embedded at existing decision points, most notably in preoperative and surgical clinics, and it must automatically translate into a small number of default, measurable actions. Briefly, this is well aligned with core implementation elements for prediction models(39). The Consolidated Framework for Implementation Research (CFIR) emphasizes that intervention characteristics, local context, and workflow compatibility determine uptake, while RE-AIM focuses evaluation on reach, effectiveness, adoption, implementation fidelity, and maintenance(40, 41). Implementation strategies with the strongest track record in acute care include iterative workflow co-design, clinical champions, EHR enabled order sets, audit and feedback, and clear ownership of escalation triggers.

Because Aims 1 and 2 involve retrospective medical record review and clinician-facing triage evaluation, patient safety risks are minimal. For Aims 3 and 4, RSR scores will be evaluated for predictive performance and clinical utility while preserving clinician oversight: during predictive validation, scores will not independently determine clinical care, and during pathway activation, RSR-informed recommendations will be implemented only within existing institutional standards and at the discretion of the treating clinical team. RSR scores are investigational and will not replace clinician judgment, alter urgent care decisions, or serve as the sole basis for delaying, denying, or approving surgery.

### Limitations

This proposal has several limitations. First, as a conceptual and protocol-oriented framework, RSR is not yet supported by prospective outcome data demonstrating that score-guided pathway activation improves patient-centered recovery; the staged validation plan is intended to address this evidence gap. Second, while the rubric is designed to be pragmatic, domain assignment will still be subject to inter-rater variability, particularly for elements that rely on clinical judgment or incomplete documentation; reliability testing and structured training will be required for scale-up. Third, the Rescue domain is deliberately context-dependent, reflecting local staffing, monitoring, and escalation capacity more than patient biology, which improves actionability but may limit transportability and require local calibration. However, the flexibility of the rubric allows clinicians to incorporate their experience with their healthcare system into perioperative decision-making. Fourth, any triage tool risks unintended consequences, including over-triage (resource strain, unnecessary ICU utilization), under-triage, and testing cascades if score outputs are not tightly linked to management-changing bundles; implementation should therefore measure fidelity, balancing metrics, and equity to ensure that pathway activation does not amplify disparities. Finally, reducing complex perioperative trajectories to ordinal scores necessarily sacrifices granularity; RSR should be used to structure decisions and communication, not to replace clinician judgment or shared decision-making.

### Conclusions

This proposed framework and protocol reframes high risk in perioperative care as a potentially modifiable mismatch between physiologic reserve, surgical stress, and system rescue capacity, rather than a fixed patient label. By translating this interaction into a pragmatic, clinician-facing RSR triage rubric designed to trigger actionable, bundled pathways and resource-aligned postoperative planning, the framework aims to move preoperative evaluation beyond clearance toward risk mitigation, shared decision-making, and prevention of postoperative deterioration and loss of independence. The next step is staged validation of feasibility, reliability, prognostic performance, and clinical utility to determine whether RSR-guided care improves patient-centered recovery while avoiding unintended over-triage and low-yield testing cascades.

## Author Contributions

Inwoo Sohn, Tanush Singh, and Zyad J. Carr contributed to Conceptualization, Methodology, Writing, Original Draft and Writing, Review & Editing. ZJC additionally contributed to Supervision and Resources. All authors reviewed and approved the final manuscript.

## Data Availability

No datasets were generated or analysed during the current study. All relevant data from this study will be made available upon study completion.

## Acknowledgements

None

## Supporting Information

None

